# MantisCOVID: Rapid X-Ray Chest Radiograph and Mortality Rate Evaluation With Artificial Intelligence For COVID-19

**DOI:** 10.1101/2020.05.04.20090779

**Authors:** Yağmur Yaşar, Berat Tuna Karli, Cem Çöteli, Mert Burkay Çöteli

## Abstract

The novel coronavirus pandemic has negative impacts over the health, economy and well-being of the global population. This negative effect is growing with the high spreading rate of the virus. The most critical step to prevent the spreading of the virus is pre-screening and early diagnosis of the individuals. This results in quaranteeing the patients not to effect the healthy population. COVID-19 is the name of the disease caused by the novel coronavirus. It has a high infection rate and it is urgent to diagnose many patients as we can to prevent the spread of the virus at the early stage. Rapid diagnostic tools development is urgent to save lives. MantisCOVID is a cloud-based pre-diagnosis tool to be accessed from the internet. This tool delivers a rapid screening test by analyzing the X-ray Chest Radiograph scans via Artificial Intelligence (AI) and it also evaluates the mortality rate of patients with the synthesis of the patient’s history with the machine learning methods. This study reveals the methods used over the platform and evaluation of the algorithms via open datasets.

## 1 Introduction

COVID-19 is the infectious disease caused by the most recently discovered novel coronavirus. This new virus and the caused disease were unknown before the outbreak began in Wuhan, China, in December 2019. COVID-19 is now a pandemic (WHO announced at March 12) affecting many countries globally [1]. COVID-19 can appear with many symptoms such as fever, fatigue, dry cough, anorexia, myalgia (muscle ache), dyspnea(shortness of breath), expectoration (the coughing up and spitting out of material from the lungs, bronchi, and trachea, sputum), pharyngalgia, diarrhea (loose, watery stools),nausea, dizziness, headache, vomiting and abdominal pain. Respiratory system symptoms are more common, and onset of disease pneumonia appears to be the most frequent serious manifestation of infection, characterized primarily by fever, cough, dyspnea, and bilateral infiltrates on chest imaging [2][16]. For the diagnosis, researchers define gold standard and lateral methods to overwhelm the pandemic via early diagnosis, but they have disadvantages such as high duration for RT-PCR (Reverse transcription polymerase chain reaction), low sensitivity for antigen/antibody tests, high radiation exposure for Computer Tomography (CT). Since these methods have many disadvantages, a rapid diagnosis pre-screening test is crucial to keep the population well-being. Physicians’ labor is the most important factor to fight with the novel coronavirus. In addition, decision support systems (DSS) which are improved with the Artificial Intelligence (AI) and machine learning methods are timesaving activities for the physicians. A rapid analysis for the Chest X-ray (CXR) scans, CT, Infection Rate or Mortality Rate with the machine learning methods are some of the helpful tools and researchers are trying to build such tools for pre-screening COVID-19. This study defines a deployed environment^1^ for rapid evaluation of the mortality rate and CXR scans via machine learning tools. The analysis explained in this study is the submitted work for the #EuvsVirus Hackathon^2^. The article goes as follows: Sec. 2 discusses about the diagnosis methods and their disadvantages. Sec. 3 reveals the algorithms in detail. Sec. 4 is the evaluation of each algorithm deployed over the platform and Sec. 5 concludes the work.

## 2 Background

RT-PCR is the gold standard to diagnose the COVID-19 [3] but it is necessary to keep in mind that there are not enough studies to evaluate the accuracy and predictive values of the tests. Sensitivity of testing depends on the precise of RT-PCR assay, type of specimen, duration of illness at testing. In a study at China, 1070 samples collected from 205 patients. Bronchoalveolar lavage fluid had the most positive results 14 of 15 patients (93 percent). Specimens from upper respiratory tract had fewer positive results. From 326 pharyngeal swabs there were 136 positive results (32 percent) and from 8 nasal swabs there were 5 positive results (63 percent). Researchers in this study suggest that testing specimens from multiple sides may improve the sensitivity [4]. According to the statement and the study before it is necessary to do studies for evaluating RT-PCR tests.

The second diagnostic tool defined in the literature is CT. According to a study in Wuhan, China with 1014 patient’s chest CT has 97 percent sensitivity based on positive RT-PCR results. Researchers suggest that chest CT could be a primary tool to diagnose COVID-19. Unfortunately, specificity was 25 percent in other words 75 of 100 patients were false positive [5]. CT scan on pediatric patients should also be avoided due to radiation [6]. Besides that, the problem is that resources to diagnose are not sufficient. In the literature, machine learning methods are also applied for the diagnosis through CT images [18] [19] and they are used for supporting the clinician’s decisions. However, they can not be applied in large number of samples to exploit the speed of AI interpretation. Therefore, lateral and rapid diagnostic tools are required for pre-elimination steps.

According to the statement before pneumonia is the most common clinical manifestation and has fatal outcomes. That is the inspiration of solution. In order to diagnose the pneumonia, CXR appears to be a useful tool and to diagnose the community acquired pneumonia. In fact, it is a gold standard with clinical evaluation and microbiologic test results [7]. Unfortunately, for COVID-19 CXR has some limitations. In a study on 64 patients some patients had no abnormality in their CXR even they had positive initial RT-PCR. CXR may be normal in early or mild disease [8], but this test can be used more frequently due to low radiation exposure. The health systems of the governments also have enough x-ray scan devices to satisfy the population. In spite of that CXR is a low-cost diagnostic tool and in healthcare chest x-rays are used frequently and we should keep in mind that that study has limitations as researchers stated. That inspires second part of our solution. Some researchers also publish their AI architectures according to the detection of COVID-19 patients [20] [21]. By evaluating the fact that CXR is not enough to speak about certain diagnosis, a supporting parameter can be provided to the physician for the conditions where AI can not catch the condition. In this study, this parameter is defined as mortality rate that shows the risk of mortality for the person who is infected with COVID-19 disease.

There are people who are at higher risk for severe illness. Those at high-risk from COVID-19 are people 65 years and older, people who live in nursing home or long-term care facility. Also people of all ages with underlying medical conditions, particularly if not well controlled, including, people with chronic lung disease or moderate to severe asthma, serious heart conditions, immunocompromised (Many conditions can cause a person to be immunocompromised, including cancer treatment, smoking, bone marrow or organ transplantation, immune deficiencies, poorly controlled HIV or AIDS, and prolonged use of corticosteroids and other immune weakening medications), people with severe obesity (body mass index [BMI] of 40 or higher), diabetes, chronic kidney disease undergoing dialysis, liver disease [9]. There should be a risk evaluation parameter to direct these patients to other diagnostic services and identify these patients who are at the risk.

## 3 Proposed Solution for pre-screening COVID-19

### 3.1 Decision Support System

MantisCOVID is the hybrid platform proposed via this study. The evaluation platform has two outputs after screening the group of patients as the prediction about the risk in COVID-19 via CXR and the mortality rate. Two parameters can be used as a decision support system to direct the patient for the other highly sensitive tests. In the case where mantisCOVID cannot catch COVID-19 patient via AI elimination from CXR, the physician can change approaching style to the patient via evaluating the mortality rate. If the patient has no symptoms and negative test results but have a higher mortality rate in that population, it is advised that follow up the patient closely but that advise should be evaluated by clinicians. Although forecasting of mortality rate is not easy due to dependence over many conditions, we have tried to estimate the mathematical model as linear regression in terms of 7 parameters via using the open datasets over the patients [10]. The system works over the cloud as a web page and each physician can access to the system for the available computations. The system is designed to be scattered over millions of people to provide scalability via cloud. Flow diagram of the platform is seen from Fig. 1.

**Figure 1:**
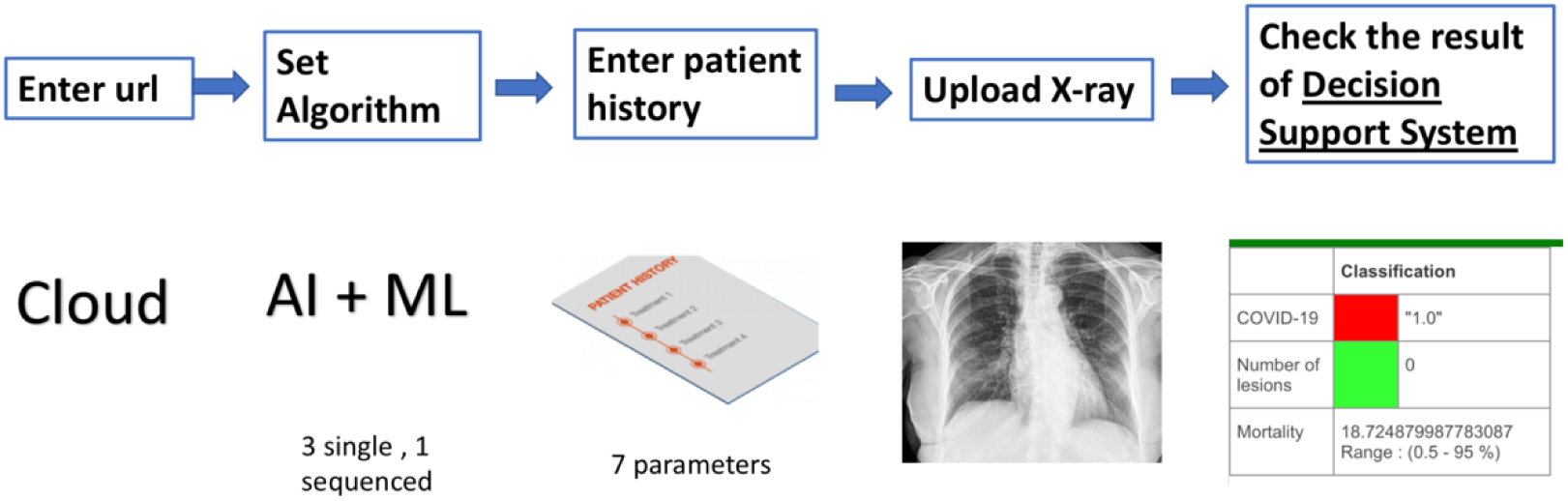
Decision Support System for Pre-Screening COVID-19

### 3.2 Methods

The system has two types of forecasting such as the mortality rate and CXR Posteroanterior (PA) chest view via AI elimination. These methods used to implement the processing flow of forecasting the mortality rate is given in the following details. During the development, open source datasets are used and the python code for data synthesis and grouping from these sets is given.^3^

The dataset used to compute the mortality rate is from open source COVID-19 Survival Calculator. The mortality rate from dataset were modeled as a linear regression method by connecting it to 7 parameters [10]. The chosen mortality rate factors were age, sex, height, weight, health conditions (asthma, carcinoma, chronic kidney disease compromised immune system, coronary heart disease chronic obstructive lung disease, diabetes, HIV positive, hypertension (high blood pressure), other chronic illness), blood type and smoking type. We have discarded some of mortality rate adjustment parameters to obtain a minimum dataset. The methods used to obtain the linear regression model for the mortality rate are the open source libraries associated with the python scikit-learn [15].

In the presentation of the CXR PA patient elimination AI platform, three types of AI algorithms are available to check whether the person is risky or not. The flow diagram can be seen from the Fig. 2. The sequence has a direct role to detect the risk from x-ray scans. As an aim, this hybrid module is not used for certain diagnosis due to low sensitivity occurring via PA Chest View. It is only used as a decision support system for the clinician.

**Figure 2:**
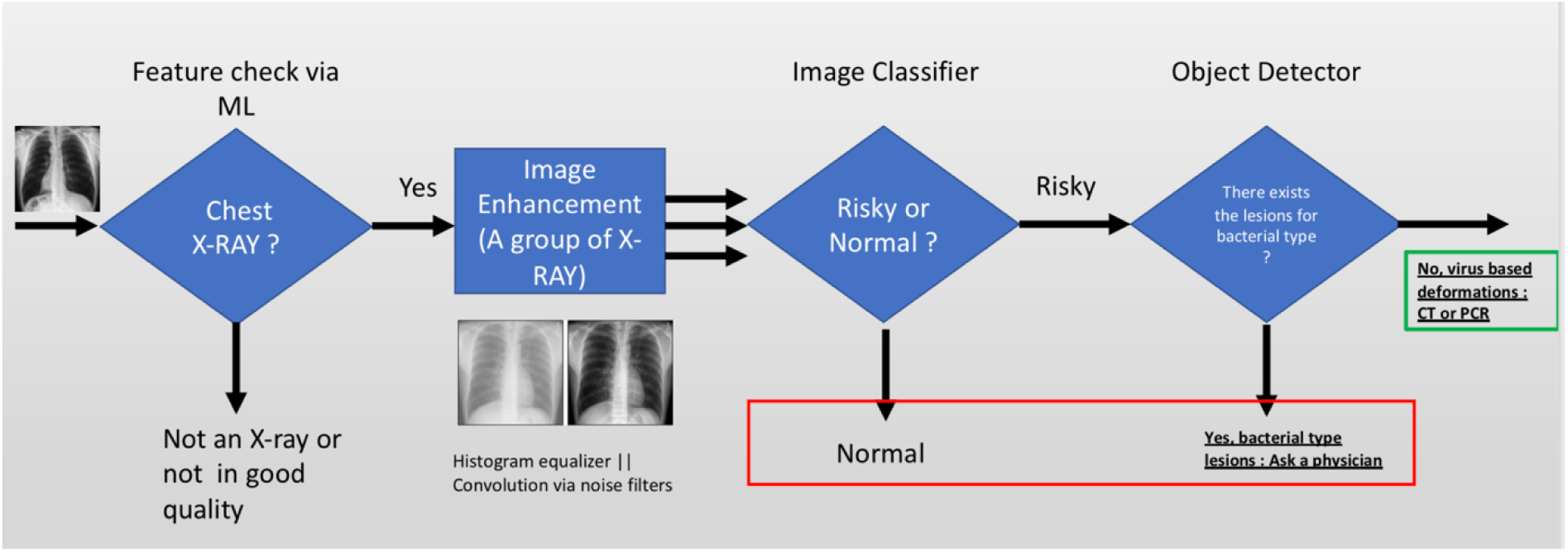
Hybrid X-Ray Analysis System for elimination of non-risky patients

The first model is generated to classify whether the input image is x-ray image or not. The model is trained via a dataset containing CXR PA scans with 211 images and images with 210 images [12] [13]. Dataset is divided into training (70 percent) and test (30 percent) sets. If an image is detected as besides of X-ray image, the other algorithms are not processed, and it warns the user to check the uploaded image.

Keras library is used to generate the Convolutional Neural Network (CNN) architecture for the starting point of the classifier [17]. In the input layer, 32 convolutional filters with size 3×3 are inserted with Rectified Linear Unit (RELU) activation function. Inputs are accepted as 200×200 with Red-Green-Blue (RGB) colors which makes input shape 200×2003. Max pooling layers and 2 hidden layers are inserted with 3×3 filter after convolution. Each hidden layer contains 32 convolutional filters with size 3×3 RELU activation functions with the addition of max-pooling layer with 2×2 filters. Then, flatten operation is applied. Fully connected layer is added with 64 units with RELU activation function. Later, dropout technique is applied with the rate of 0.5 to avoid overfitting. Finally, fully connected layer is used with a unit with sigmoid function (1, if the image is x-ray image; 0, otherwise).

The classifier AI in the middle revealing whether the patient is risky (COVID-19 / pneumonia) or not is the trained model (modified version of AlexNet [11]) according to the open datasets strengthened with the currently obtained COVID-19 patients’ day by day [12]. The dataset is split into train (0.7), test (0.2) and validation (0.1) to enable the platform for the evaluation. It totally contains 8851 normal, 121 COVID-19 and 9579 pneumonia patient data [13]. The AI Architecture of the object classifier and training batch sizes are given in Fig. 3.

**Figure 3:**
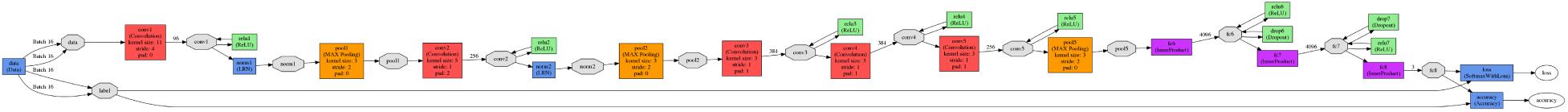
Deep Neural Network architecture offered for 3 types of CXR PA patient

The last part defined as the object detection case is the detection of the lung opacity parts associated with the bacteria type pneumonia. The AI neural network is the trained tiny version of You Only Look Once (YOLO) [14] to mark the single objects in CXR PA images. The anomaly is named as lung opacity. The dataset used in this part is the annotated CXR pA obtained through the open source datasets [13].

## 4 Evaluation

The system presented in this paper contains 4 different machine learning techniques to provide a rapid analysis of CXR PA view and the mortality rate evaluation. These algorithms are evaluated one by one with the allocated data. Each evaluation results can be seen within the following details.

1. **Linear Regression model to forecast the mortality rate:** This evaluation contains allocated 10000 patient data gathered from the open source dataset. The mean squared error and the standard deviation between the actual and predicted values are computed as 2.87 and 1.69 respectively. Fig. 4 shows the details about the actual and predicted values via testing with 150 conditions.
2. **Binary classifier to check whether uploaded image is X-ray scan or not:** This evaluation contains additional 1000 X-ray and 1030 random images to be used in the evaluation of the binary classifier algorithm. The correctly predicted values for the X-ray scans and random samples are given as 971 and 943 separately. It is observed that the averaged accuracy is about 96 percent.
3. **Classifier to separate COVID-19/Pneumonia/normal patients:** This part contains randomly selected 150 patient data to be separated via the AI algorithm. The sensitivity of the risky patient detection (COVID-19/pneumonia) is 84 percent. The sensitivity for the detection of normal patients is 67 percent. The confusion matrix can be seen in the following Fig. 4. As the main objective is the detection of risky patients in high accuracy, the system conditions can be improved via gathering more patient data which is known as COVID-19 certainly.
4. **Lesion detection over the bacteria type x-ray scans:** This part contains randomly selected 30 patient data to be analyzed via the AI algorithm. The evaluation criterion is comparing the number of lesions between the real annotated and predicted X-Ray scans. The allocated dataset is divided into three groups as 2 lesions (10 image), 1 lesion (10 image) and no lesion (10 image). The detected and the actual number of lesions over the groups are given as in the following 17/20, 9 /10 and 4/0 respectively. In fact, the algorithm determined 4 lesions over the zero annotated 10 images. The algorithm findings over the groups corresponding to 1-2 annotations match with the real annotated regions. The overall sensitivity for the lesion detection is revealed as 86 percent. The samples taken from the original, annotated and algorithm outputs are given in Fig. 5, 6, 7.

**Figure 4:**
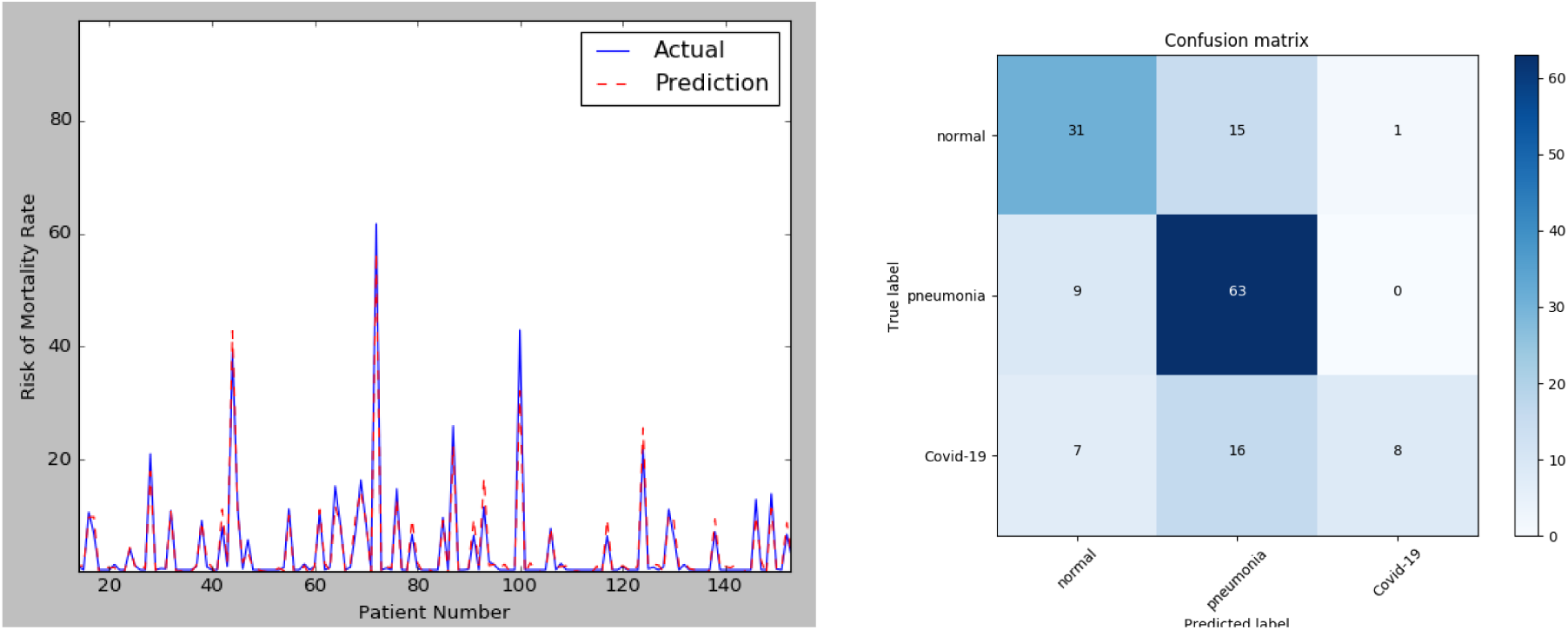
Left: Actual and prediction values of mortality rate, Right: Confusion Matrix for the classifier

**Figure 7:**
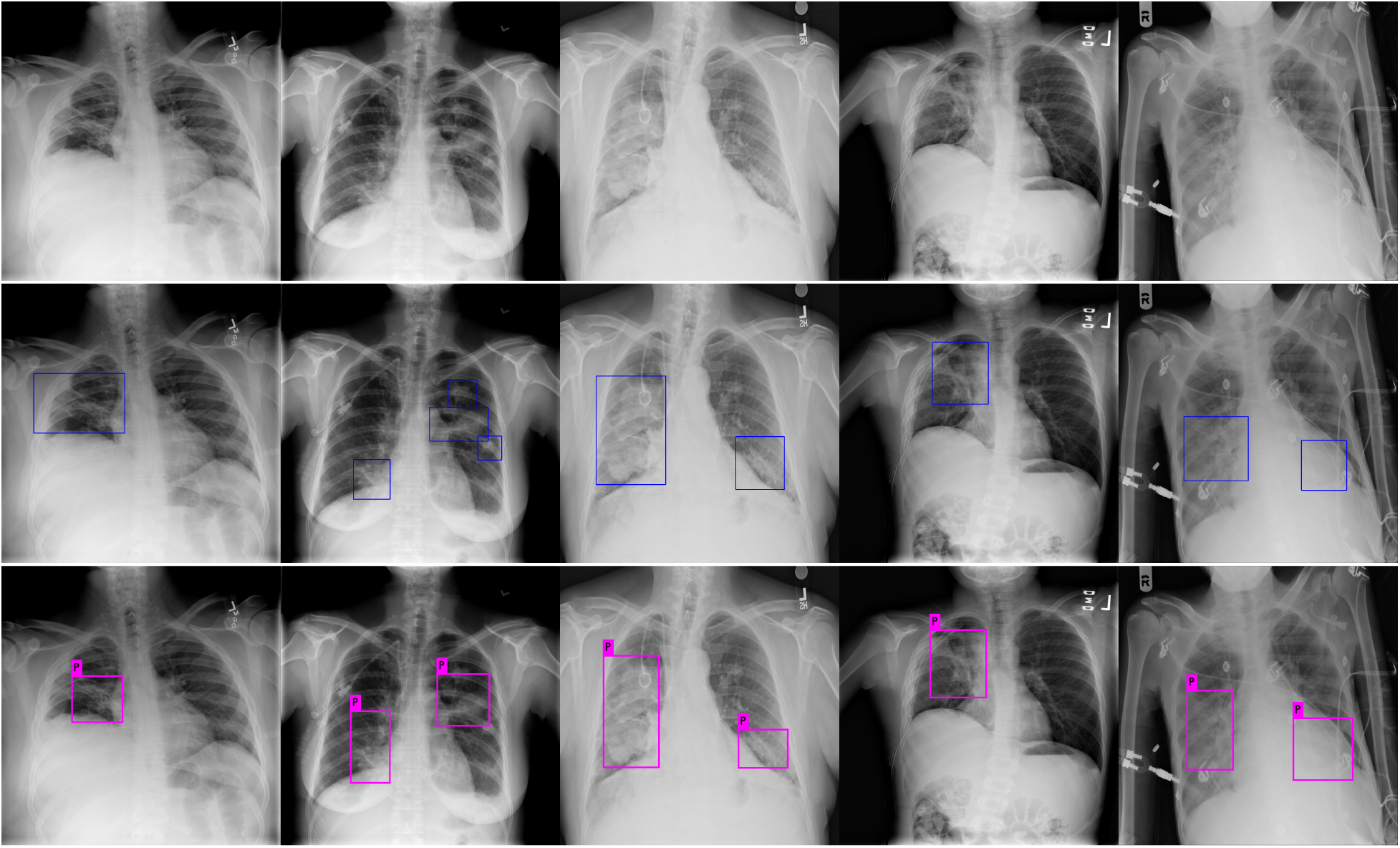
Lesion Detection Algorithm Results. From top to bottom, original CXR PA images, annotated images via experts, AI examination results respectively.

## 5 Conclusion and Future Work

MantisCOVID is a cloud-based pre-screening platform that would be useful for the elimination of non-risky patients in the coronavirus pandemic. Since the algorithms are developed and evaluated using the open source data and platforms, this system needs to be evaluated via the clinical perspective by the legal authorities. This platform is an internet-based pre-diagnosis tool to be verified via the universities during the COVID-19 diagnosis. However, a physician would also use this tool as an assistant to scan and diagnose the patients due to the rapid diagnosis requirement to prevent spreading of the coronavirus. In fact, the gold standards used to diagnose the COVID-19 patients also have high false alarm rates because of the epidemiology of the disease is not fully known.

As a future work, the clinical validation of the internet-based platform will start with the known legal authorities and other state of the art AI architectures will be used to increase the sensitivity and specificity of the platform. The integration over the platform will continue with the Chest Anteroposterior (AP) views, Chest CT and Ultrasound images by providing novel AI architectures. Infection rate for the novel coronavirus would also be evaluated with the usage of tracking the patient’s locations. After the coronavirus pandemic, this platform would also be used for scanning the society in terms of the mortality rate and X-ray scans in any other illnesses and symptoms. There should be more studies to identify case fatality rate and infection rate in the case where proper data will be collected to train AI.

## Data Availability

Dataset used in this study is open and can be accessible from the internet.

https://github.com/ieee8023/covid-chestxray-dataset

https://www.kaggle.com/c/rsna-pneumonia-detection-challenge

https://www.covid19survivalcalculator.com/calculator

## Footnotes

1 mantisCOVID: https://scan.mantiscope.com/?lang=en

2 https://euvsvirus.org/

3 https://github.com/mbcoteli/MantisCOVID

